# Alzheimer’s Polygenic Risk Scores Are Not Interchangeable: Evidence from 1,752 Models

**DOI:** 10.64898/2026.09.02.26362060

**Authors:** Elizabeth L. Ward, Peter T. Nelson, Yuriko Katsumata, David W. Fardo, Gregory A. Jicha, the Alzheimer’s Disease Neuroimaging Initiative, Justin B. Miller

## Abstract

**Introduction:** Polygenic risk scores (PRS) may improve Alzheimer’s disease (AD) risk prediction before symptom onset, yet choosing an appropriate model can be challenging.

**Methods:** Using the standardized GenoPred pipeline, 1,752 PRS models (9 algorithms; 584 configurations; 3 genome-wide association studies) were evaluated and stratified by genetic ancestry and *APOE* diplotype. PRS models were evaluated using 11,200 clinical or autopsy-confirmed AD cases and 19,321 controls age ≥65 from the Alzheimer’s Disease Sequencing Project Release 5.

**Results:** PRS results were not consistent across methodologies (Spearman’s ρ: –0.49 to 1), with >95% of individuals having PRS in both the top and bottom risk deciles. Top-performing PRS were effective at stratifying AD risk across ancestries (AFR: *P*=2.04×10^-26^; AMR: *P*=4.57×10^-21^; EAS: *P*=6.21×10^-40^; EUR: *P*=7.90×10^-187^).

**Discussion:** PRS parameters should be optimized for each ancestry. Contradictory signals across methodologies underscore the need for carefully choosing suitable PRS methods and fine-tuning algorithmic parameters to ensure accuracy and consistency.

**Data Availability:** Access to the ADSP is controlled by The National Institute on Aging Genetics of Alzheimer’s Disease (NIAGADS). All scripts used to analyze the data are freely available at https://github.com/jmillerlab/prs_comparisons.

## Introduction

The genetic, pathological, and clinical presentation of Alzheimer’s disease (AD) is highly heterogeneous and influenced by many genetic and environmental factors that make it difficult to predict disease risk *a priori*. Current estimates indicate that 60-80% of AD risk is heritable [1]. At least 75 loci are implicated by genome-wide association studies (GWAS; *P* < 5 × 10^-8^) [2] and can be used to model AD risk [3–10].

As the field moves toward developing clinical AD genetic risk models, it is important to recognize that other diseases now leverage polygenic risk scores (PRS) to prioritize follow-up testing based on genetics [11–14], influence within-patient clinical interventions [15, 16], identify genetic overlap between traits [17, 18], classify disease subtypes [19, 20], characterize genetic ancestry [21], and determine causal genetic relationships through Mendelian randomization [22–24].

PRS have been shown to be helpful in various clinical contexts, although this has not yet penetrated into dementia clinical care. PRS can identify more clinically significant incidences of prostate cancer than standard screening techniques [25] and reduce the 10-year incidence rate of major cardiovascular events [26]. These precedents indicate that appropriate PRS disclosure for AD/dementia may similarly have a positive impact on patient care in this common clinical scenario.

Genetic screening can already help indicate the extent to which modifiable risk factors (e.g., lifestyle choices and environment) can lower dementia risk. While some lifestyle choices are beneficial for all individuals, regardless of genetic predisposition [27], the most significant changes in cognition occur in individuals with low- and intermediate genetic risk, and lifestyle changes have a very limited impact on dementia risk in individuals with the highest genetic risk profiles [28]. Similarly, disease progression is faster in individuals with high PRS; a small cohort (122 individuals) with mild cognitive impairment (MCI) were significantly more likely to convert to AD within 36 months if their PRS was in the highest decile (∼94%) compared to the lowest decile (25%) [29]. As we move toward patient-centered dementia care, future clinical treatment plans and recommendations for end-of-life planning might include patient genetic risk profiles, necessitating accurate and robust comparisons of current genetic risk models to ensure that appropriate methods are used to predict AD risk for each population.

To date, benchmarking studies generally use relatively small cohorts to assess various methodologies or larger cohorts to assess fewer models. Bellou, Kim [30] showed that GWAS summary statistics, more than algorithmic choice, account for much of the differences in prediction accuracy [30], yet it is difficult to generalize those results given the relatively small sample sizes of their test sets, which included the European population from the Alzheimer’s Disease Neuroimaging Initiative (ADNI; *n*=568) and BioFINDER (*n*=766). Other benchmarking efforts have focused on computing the optimal threshold for p-value clumping [31], and improving the transferability of PRS to individuals from non-European ancestries such as Korean [32] and Hispanic [33] populations. Yet, PRS can often be unreliable at the individual-level [34], making single risk estimates inconsistent.

Various issues with PRS for AD are well-documented [35] and include challenges with replicability and interpretability, high variance between different populations, contradictory signals from different GWAS, and incongruence between PRS derived from different methodologies. Kullo [36] outlined current barriers to clinical PRS implementation, suggesting that PRS should be considered one facet of a multifactorial framework for risk stratification. As such, PRS are often incorporated in large models that consider PRS influence in conjunction with other covariates [37].

Here, we performed the most extensive comparative analysis of AD PRS to date. We compared nine distinct PRS algorithms with 584 hyperparameter configurations (1,752 total PRS) for each of the three most commonly used AD GWAS (Kunkle, Grenier-Boley [38], Jansen, Savage [39], and Bellenguez, Küçükali [2]) using 11,200 clinical or autopsy-confirmed AD cases and 19,321 controls age ≥65 from the latest Alzheimer’s Disease Sequencing Project (ADSP) Release 5 (R5). PRS were stratified by genetically inferred ancestry and *APOE* diplotype, and we examined how algorithmic, hyperparameter, and GWAS choice impact PRS results. We anticipate that these benchmarking results will help shape the methodological choice and framework for future research and clinical PRS utilization.

## Materials and Methods

### Overview

A brief overview of the workflow for generating and analyzing AD PRS across various genetic ancestries in the ADSP R5 (Repository site: dss.niagads.org; release: NG00067.20; Preview Joint Called VCF) whole-genome sequencing dataset is shown in **Figure 1**. The pipeline includes preprocessing of the target cohort and three large-scale GWAS summary statistics, followed by PRS calculation using the GenoPred pipeline (3 GWAS x 9 algorithms with hyperparameter tuning, yielding 1,752 PRS per genetic ancestry). Downstream analyses were stratified by genetically inferred ancestries and encompass correlation-based selection of uncorrelated PRS, evaluation of PRS consistency and variance at the individual level, performance assessment for each *APOE* diplotype, and an assessment on how potential sample overlap with the original GWAS might affect the results. Samples were required to have matching *APOE* diplotyping calls through *APOE* diplotyping and whole-genome sequencing (WGS). Samples with contradictory *APOE* diplotypes or missing data were removed from downstream analyses.

**Figure 1.**
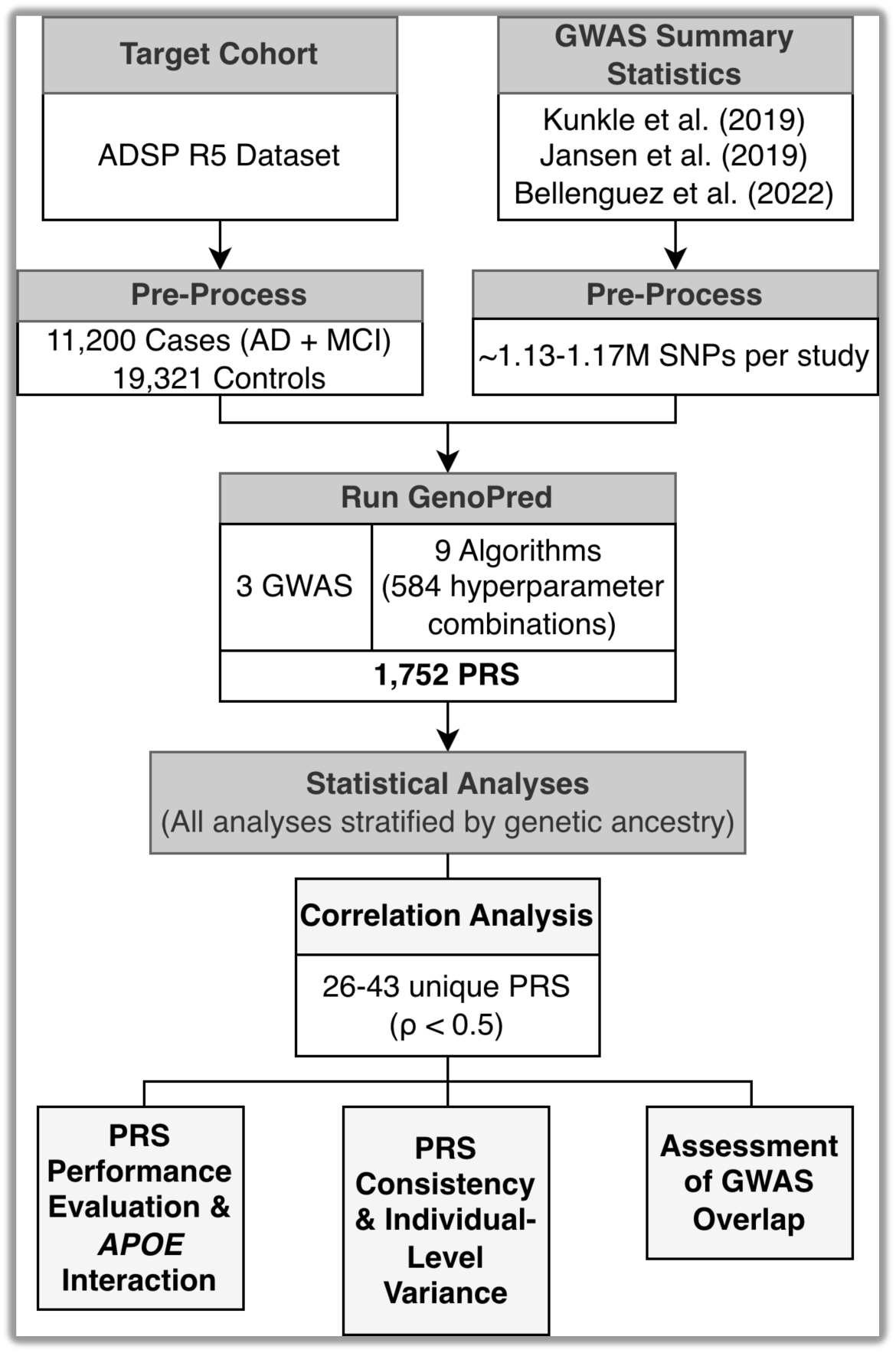
Overview of the analytical pipeline for PRS computation and evaluation using the ADSP R5 cohort.

### GWAS summary statistics

We utilized GWAS summary statistics from the three most commonly-used large-scale meta-analyses of AD risk, all primarily from European-ancestry individuals. Kunkle, Grenier-Boley [38] performed a meta-analysis across the Alzheimer’s Disease Genetic Consortium (ADGC), the European Alzheimer’s Disease Initiative (EADI), the Cohorts for Heart and Aging Research in Genomic Epidemiology (CHARGE), and the Genetic and Environmental Risk in AD/Defining Genetic, Polygenic and Environmental Risk for Alzheimer’s Disease Consortium (GERAD/PERADES). In total, the meta-analysis included 35,274 clinically diagnosed or autopsy-confirmed AD cases and 59,163 controls (total *n*=94,437), identifying new risk loci implicated in Aβ, tau, immunity, and lipid processing pathways.

Jansen, Savage [39] performed a GWAS-by-proxy (GWAX) by integrating clinically diagnosed AD with AD-by-proxy phenotypes (parental dementia history; r=0.81 correlation with clinical AD), encompassing 71,880 cases and 383,378 controls (total *n*=455,258) from the Psychiatric Genomics Consortium (PGC-ALZ), the whole-exome sequencing (WES) data from ADSP (4,343 cases and 3,163 controls), the International Genomics of Alzheimer’s Project (IGAP), and the UK Biobank (UKB), revealing 29 distinct risk loci enriched in immune tissues and amyloid degradation.

Bellenguez, Küçükali [2] conducted a two-stage meta-analysis via the European Alzheimer and Dementia Biobank (EADB) consortium, totaling 111,326 AD cases (both clinically diagnosed and proxy) and 677,663 controls (total *n*=788,989), uncovering 75 risk loci (42 novel) associated with amyloid/tau pathways, lipid metabolism, endocytosis, and microglial function. **Supplementary Table S1** describes key characteristics of the three GWAS/GWAX employed in our PRS calculations.

### Target cohorts and sample selection

This study uses 57 harmonized cohorts from the ADSP R5. We extracted sample IDs from the ADSP with a confirmed diagnosis of Control, MCI, or AD. We excluded individuals <65 years old or lacking annotation for *APOE* diplotype, age, sex, or race. Additional exclusions included individuals with non-AD diagnoses: Braak stages 1-5 without an AD diagnosis (*n*=968), frontotemporal dementia (*n*=169), Lewy body dementia (*n*=206), machine learning-called cases/controls (*n*=1,008), Parkinson’s disease (*n*=11), pathological aging (*n*=39), progressive supranuclear palsy (*n*=16), vascular dementia (*n*=266), or other dementia cases (*n*=6,426).

We then compared the sample IDs from the data dictionaries with the sample IDs from the variant call format (VCF) files, identifying 25 mismatches (**Supplementary Table S2.1**). After review, 21 IDs were updated to align with prior releases, and the remaining four individuals were excluded (**Supplementary Table S2.2**).

Given that MCI is a high-risk transitional stage of dementia with substantial conversion rates to AD within a few years [40, 41], we merged the MCI and AD groups for all downstream analyses.

Some data were obtained from the Alzheimer’s Disease Neuroimaging Initiative (ADNI) database (adni.loni.usc.edu), as ADNI participants are included among the cohorts sequenced and harmonized within the ADSP. The ADNI was launched in 2003 as a public-private partnership, led by Principal Investigator Michael W. Weiner, MD. The primary goal of ADNI has been to test whether serial magnetic resonance imaging (MRI), positron emission tomography (PET), other biological markers, and clinical and neuropsychological assessment can be combined to measure the progression of MCI and early AD.

### Preprocessing and quality control (QC) of the target cohort

We used the standardized GenoPred pipeline [42] to perform target genotype QC, relatedness assessment, ancestry inference, GWAS summary statistics harmonization, PRS calculation, and score standardization. GenoPred leverages the 1000 Genomes Project Phase 3 + Human Genome Diversity Project (1KG+HGDP) reference panels restricted to HapMap3 variants for ancestry inference and (by default) uses European LD references as proxy for non-European ancestries.

### Relatedness assessment

We used GenoPred to computationally infer four genetic ancestries: Admixed American (AMR), African (AFR), East Asian (EAS), and European (EUR). GenoPred assesses relatedness using the KING estimator [43] implemented in PLINK v1.9 on the full target sample. The KING estimator provides a robust measure of pairwise kinship coefficients by quantifying allele sharing patterns across variants in a way that is largely insensitive to population structure or substructure in the sample. Individuals with kinship coefficient >0.044 (fourth-degree relatives or closer) were removed to minimize bias and excluded from all downstream analyses.

### Ancestry assignment and within-population QC

Genetic ancestry was assigned using a multinomial elastic-net model trained on the 1KG+HGDP reference panel. The first 6 principal components (PCs) were projected into the target data using overlapping HapMap3 variants (minor allele frequency [MAF] > 0.05, missingness < 0.02, Hardy-Weinberg equilibrium [HWE] *P* > 1 × 10^-6^, LD-pruned: window = 1000kb, step = 5, r^2^ < 0.2). Samples were classified into each ancestral population only if the maximum posterior probability was >95%.

Within-population QC followed GenoPred’s standard procedures. For populations with ≥100 individuals, ten within-population PCs were computed on unrelated individuals using pruned variants, and outliers were excluded based on k-means centroids. Individuals with self-reported Native Hawaiian (*n*=11) or American Indian (*n*=78) race were excluded from concordance analyses due to low numbers and lack of matched references. Due to insufficient sample sizes post-filtering, Central/South Asian (CSA; *n*=39) and Middle Eastern (MID; *n*=9) groups were excluded from PRS analyses (see **Supplementary Table S3** for demographic characteristics of excluded ancestral populations).

### Target cohort composition and demographics

After all filtering, downstream PRS analyses were stratified by four ancestral populations with adequate sample sizes: AFR (*n*=3,393; 1,260 cases; 2,133 controls), AMR (*n*=3,212; 1,304 cases; 1,908 controls), EAS (*n*=2,289; 1,018 cases; 1,271 controls), and EUR (*n*=13,232; 4,392 cases; 8,840 controls). Demographics of these groups are presented in **Table 1**.

**Table 1.** Demographics of the final target cohort.

| Ancestry | N | | Age, mean ± SD | | Female, % | | APOE $\epsilon 4$ Carriers, % | |
| --- | --- | --- | --- | --- | --- | --- | --- | --- |
|  | Cases | Controls | Cases | Controls | Cases | Controls | Cases | Controls |
| Admixed American | 1,304 | 1,908 | 75.98 ± 6.55 | 74.09 ± 6.42 | 64.80 | 67.35 | 32.82 | 20.49 |
| African | 1,260 | 2,133 | 76.70 ± 6.77 | 76.20 ± 7.20 | 72.06 | 77.26 | 57.46 | 35.26 |
| East Asian | 1,018 | 1,271 | 75.39 ± 5.65 | 76.57 ± 4.94 | 60.81 | 48.62 | 53.14 | 27.85 |
| European | 4,392 | 8,840 | 76.41 ± 7.65 | 78.61 ± 7.46 | 54.12 | 59.08 | 57.13 | 33.04 |

### Sample- and variant-level QC

We applied no additional genotype filters beyond those used by the ADSP: FILTER=’PASS’, VQSR tranche ≥99.7%, DP ≥10, GQ ≥20, call rate>80%, mean depth<500, monomorphic variants excluded, Mendelian inconsistencies removed.

We restricted PRS calculations to HapMap3 variants, which is commonly used to improve computational efficiency without sacrificing accuracy [44, 45]. We also merged GWAS summary statistics by their reported reference genome, flipped variants to match the reference, and filtered the GWAS data to remove variants with MAF < 0.01, duplicate IDS, out-of-bounds *p-*values, or invariant sites. After filtering, the following variant counts were retained for each GWAS: Bellenguez, Küçükali [2] (1,160,429 variants), Jansen, Savage [39] (1,129,967 variants), and Kunkle, Grenier-Boley [38] (1,174,506 variants).

To align with GenoPred assumptions, which require no half-calls or multiallelic sites, we used bcftools [46] to pre-filtered ADSP VCFs to include only HapMap3 variants (`bcftools view ––regions-file)`, split multiallelic variants (`bcftools norm -m any`), and set half-calls to missing (using `bcftools view` and `awk`). We did not impute variants since we used whole-genome sequencing data and to avoid potential imputation bias, given high HapMap3 retention in our target samples. The sample-level QC threshold was relaxed to ≥60% HapMap3 variants present per individual (versus default ≥70%), which resulted in all target samples being included in these analyses.

### Polygenic risk score (PRS) calculation

We evaluated AD PRS methodologies using three GWAS summary statistics and nine algorithms standardized with GenoPred (**Table 2**). Hyperparameter tuning was performed systematically, yielding 584 PRS per GWAS (1,752 total).

**Table 2.** PRS algorithms used in comparisons.

| Algorithm | Type of Algorithm | Primary Hyperparameter(s) Tuned | Key Optimization | Number of Models Evaluated |
| --- | --- | --- | --- | --- |
| DBSLMM [47] | Deterministic Bayesian sparse LMM | None (deterministic model) | 1-28x faster, 75-93% less memory than MCMC methods | 3 |
| Lassosum [48] | Penalized regression (LASSO/EN) | $\lambda$ (shrinkage), $s$ (elastic-net mixing) | $\sim 10x$ faster than LDpred via LASSO approximation | 80 |
| Lassosum2 [49] | Penalized regression (LASSO/EN) | $\lambda$, $s$ | Improved LD stability in sparse traits; efficient parallel use | 120 |
| LDpred2 [50] | Bayesian regression (point-normal prior) | Proportion causal ($p$), heritability ($h^2$), sparsity mode | 5-100x faster than PRS-CS; handles long-range LD | 128 |
| MegaPRS [51] | Bayesian multiple regression (mixture priors) | Choice of prior (4 options) | Flexible priors | 236 |
| PRS-CS [44] | Bayesian regression (continuous shrinkage prior) | $\phi$ (global shrinkage) | Robust across heritability; strong for polygenic/sparse traits | 5 |
| PLINK pT+clump [52] | Clumping + P-value thresholding | P-value cutoff | Extremely fast and simple for large datasets | 10 |
| QuickPRS [51] | Bayesian multiple regression (optimized mixtures) | Choice of prior (inherits MegaPRS) | 10-20x faster than MegaPRS | 1 |
| SBayesRC [53] | Bayesian regression w/ annotation-informed mixtures | Mixture components, annotation sets | Uses annotations; 10-20% better cross-ancestry prediction | 1 |

### Statistical Analyses

#### Correlation-based selection of uncorrelated PRS

To determine the extent to which different GWAS summary statistics, algorithms, and hyperparameter tuning provide independent information, we evaluated pairwise correlations among all 1,752 PRS (ignoring AD diagnosis) using Spearman’s rho (ρ). Correlations were computed separately within each genetic ancestry group.

We applied a greedy selection algorithm to identify a reduced set of largely independent PRS: at multiple Spearman’s ρ thresholds (|ρ| > 0.1, 0.3, 0.5, 0.7, 0.9, or =1), clusters of highly correlated PRS were identified, and the PRS with the lowest mean absolute correlation to all others in the cluster was retained as the representative PRS. This process was repeated iteratively until no further clusters exceeded the threshold. Pairwise correlation and greedy selection analyses (**Figure 2**) were performed across all ancestry groups.

**Figure 2.**
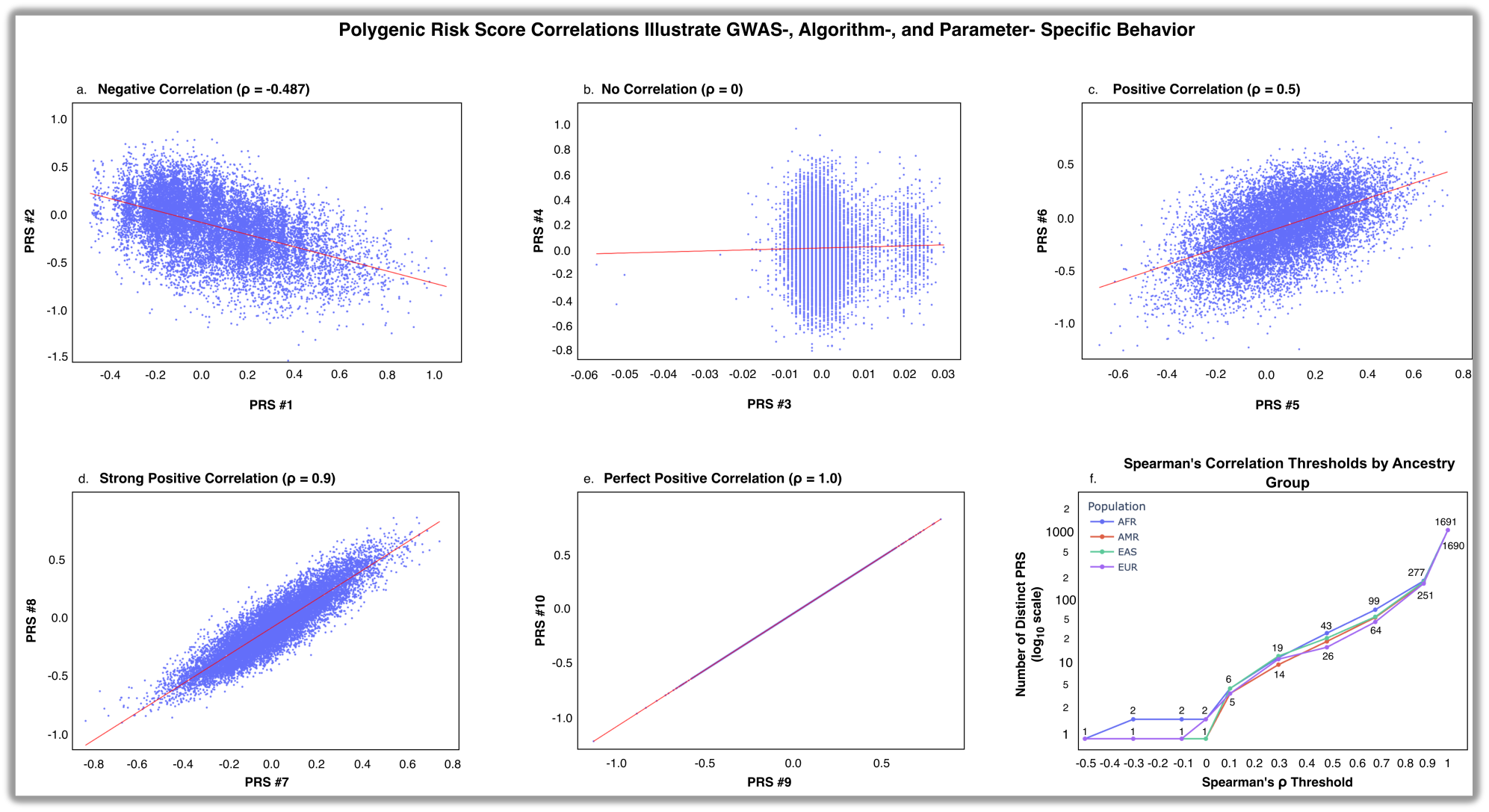
Correlations between PRS illustrate the influence of GWAS source, algorithm, and modeling parameters. Pairwise scatterplots display correlations between standardized PRS calculated in EUR ancestry participants from the ADSP cohort. Each blue point represents one individual; red lines show linear regression fits. Spearman’s rank correlation coefficient (ρ) is displayed in each panel title. Panels (a-e) highlight a spectrum of correlation strengths between different PRS models. Full model specifications (e.g., GWAS sources, algorithms, and hyperparameters) are included in **Supplementary Table S15**. Panel f shows the number of distinct PRS retained across ancestry groups after greedy feature selection, plotted as a function of the maximum Spearman’s ρ threshold.

#### PRS evaluation across *APOE* diplotypes and ancestries

We evaluated how PRS stratify AD risk across ancestries and interact with the *APOE* diplotype. We first identified ancestry-specific PRS models and thresholds that optimized separation of AD cases. Within each ancestry-specific group, we evaluated a set of largely independent PRS models (Spearman’s ρ <0.5) and selected the best PRS and corresponding threshold that maximized positive predictive value, subject to the constraint that at least 20% of individuals in that ancestry exceeded the threshold. This approach aimed to identify thresholds capturing a broadly applicable increase in AD prevalence between the elevated-risk tail and the remainder of the distribution, rather than extreme risk.

We then characterized the continuous relationship between PRS and AD prevalence across the full spectrum of values using the selected PRS models that maximized precision for the average use case. The percentage of individuals with AD was plotted as a function of standardized PRS, stratified by genetic ancestry and *APOE* diplotype.

To further illustrate the extent to which *APOE* modifies the risk differences at the chosen cutoff, we plotted the proportion of individuals with an AD diagnosis below and above the selected thresholds, stratified by ancestry and *APOE* diplotype. Differences in AD prevalence above versus below thresholds were assessed using chi-square tests.

#### Evaluating the Linearity of PRS-AD Risk Association

To test whether the association between PRS and AD risk differs above versus below the identified threshold, logistic regression models were fit separately within each ancestry-stratified subgroup using binomial logistic regression (*glm*, R verion 4.6). Each subgroup was analyzed using the PRS model and threshold that maximized precision for that ancestry. The outcome was binary AD case-control status. The primary model included continuous PRS, a binary above-threshold indicator, interaction term, age, sex, and *APOE* diplotype as covariates, with ε3/ε3 as the reference category. The interaction term directly tests whether the slope of PRS on log-odds of AD risk differs significantly between the two segments defined by the threshold. A likelihood ratio test (LRT) comparing the interaction model to a nested main-effects model (without the interaction term) assessed whether the interaction significantly improved model fit. Odds ratios (ORs) and 95% confidence intervals (CIs) were obtained by exponentiating model coefficients. The slope below the threshold corresponds to the exponentiated PRS coefficient. The slope above the threshold was computed as the sum of the PRS and interaction coefficients on the log-odds scale, then exponentiated.

#### Assessment of potential GWAS sample overlap bias

Importantly, PRS assessments assume that the target data are independent from the base samples used in the original GWAS [54]. We evaluated potential biases arising from sample overlap between the ADSP R5 WGS dataset and samples used to calculate the base GWAS summary statistics. Although we expected overlap-related biases to occur primarily among EUR-ancestry participants since all three base GWAS were restricted to EUR individuals, we also extended these assessments to AFR and AMR populations. This extension was motivated by our observation that more than 39% of individuals in each of these non-EUR groups had either prior WES data from an earlier ADSP release or originated from Alzheimer’s Disease Genetics Consortium (ADGC) cohorts that contributed samples to the Bellenguez, Küçükali [2] GWAS (**Supplementary Table S4**).

To assess potential overlap with the Kunkle, Grenier-Boley [38] and Jansen, Savage [39] GWAS (both of which included WES from an earlier ADSP release), we stratified ADSP participants by sequencing type. We classified participants as “likely overlapped” if they had both WGS and prior WES data, and as “unlikely overlapped” if they had WGS data only. We then compared PRS distributions between these two groups within the EUR, AFR, and AMR populations.

For the Bellenguez, Küçükali [2] GWAS (which included 17,141 WGS samples from ADGC cohorts), we took a conservative approach. We compared PRS distributions from participants from ADGC-affiliated cohorts (likely overlapped) and those from non-ADGC sources (unlikely overlapped) in the EUR, AMR, and AFR populations. **Supplementary Table S5** includes a list of the cohorts included within the ADGC.

We evaluated predictive performance of the top-performing PRS models in the full ADSP R5 cohort and in subgroups after excluding likely-overlapped individuals. We also performed detailed PRS distribution comparisons between likely-overlapped and non-overlapped groups using Mann-Whitney U and Kolmogorov-Smirnov tests. We applied Bonferroni correction separately within each family of tests to account for multiple comparisons. For the 1,168 PRS comparisons (WES vs. no-WES) involving the Kunkle, Grenier-Boley [38] and Jansen, Savage [39] GWAS, the corrected significance threshold was α = 0.05 / 1168 = 4.28 × 10^-5^. For the 584 PRS comparisons (ADGC vs. non-ADGC) involving the Bellenguez, Küçükali [2] GWAS, the corrected significance threshold was α = 0.05 / 584 = 8.56 × 10^-5^. Results of PRS distribution comparisons and performance metrics in non-overlapped EUR subgroups are reported in **Supplementary Tables S6-S13**. Since including or excluding the potentially overlapping samples did not result in significant differences in the evaluative metrics, we report all primary PRS analyses—including performance metrics, benchmarking, and selection of top-performing models—using the full ADSP R5 cohort.

## Results

### Genetic ancestry concordance with self-reported race

We found that only ∼65% of self-reported or administratively assigned race annotations reported in the ADSP correspond with the most common genetically-inferred ancestry for that group (**Table 3**). In the ‘White’ reported race category, which is often used as a proxy for EUR ancestry, only 70.9% of people had genetic ancestry with ≥95% probability of mapping to the EUR genetic ancestry; ‘Black’ was only slightly better with 74.7% concordance with the AFR ancestry. The ‘Asian’ race, on the other hand, strongly aligned with the EAS genetic ancestry (96.4% concordance), while the ‘Other’ race was the most genetically diverse (only 30% concordance with AMR).

**Table 3.** Concordance of reported race to genetically inferred ancestry. ^1^An additional 34 individuals reported Central/South Asian ancestry; inclusion of those individuals yields 97.9% for the Asian group overall. ^2^More individuals fell into the “No Projection” category (n=3,457) than Admixed American, but “No Projection” does not represent a single genetic ancestry and was thus excluded from this table.

| Reported Race | Most Similar Genetic Ancestry | Number of People in Reported Race Group | Number of People with Same Genetic Ancestry (Percent of Genetic Ancestry Group) |
| --- | --- | --- | --- |
| Asian | East Asian | 2,364<sup>1</sup> | 2,280 (96.4%) |
| Black | African | 4,335 | 3,238 (74.7%) |
| White | European | 18,543 | 13,147 (70.9%) |
| Other | Admixed American<sup>2</sup> | 5,193 | 1,556 (30.0%) |

Although reported race is not often a good proxy for genetic ancestry, genetic ancestry is an excellent proxy for reported race for EUR (99.4% concordance with ‘White’), EAS (99.6% concordance with ‘Asian’), and AFR (95.4% concordance with ‘Black’) individuals (**Table 4**). In contrast, concordance between genetic ancestry and race was poor in AMR (48.4% concordance with ‘Other’), CSA (75.6% concordance with ‘Asian’), and MID (55.6% concordance with ‘White’), potentially due to limited options for reporting race in the ADSP dataset. The “No Projection” group (*n*=8,345) most commonly corresponded with the ‘White’ race (45.7%).

**Table 4.** Concordance from genetic ancestry to most common reported race. Note: “No Projection” indicates insufficient evidence to assign genetic ancestry with >95% probability.

| Genetic Ancestry | Most Commonly Reported Race | Number of People in Ancestry Group | Number of People in Most Commonly Reported Race (Percent of Genetic Ancestry Group) |
| --- | --- | --- | --- |
| Admixed American | Other | 3,212 | 1,556 (48.4%) |
| African | Black | 3,393 | 3,238 (95.4%) |
| Central and South Asian | Asian | 45 | 34 (75.6%) |
| East Asian | Asian | 2,289 | 2,280 (99.6%) |
| European | White | 13,232 | 13,147 (99.4%) |
| Middle Eastern | White | 9 | 5 (55.6%) |
| No Projection | White | 8,345 | 3,811 (45.7%) |

### PRS correlation analysis

We computed 1,752 PRS using summary statistics from three GWAS combined with nine algorithms and extensive hyperparameter tuning. After excluding 40 invariant scores in each population (and 48 in the MID population), a pairwise Spearman’s rank correlation analysis was conducted between each PRS, independent of diagnostic status.

PRS results were not consistent across methodologies (**Figure 2**). Correlation between methods varied widely, ranging from moderately negative (ρ=-0.487; **Figure 2 panel a**) between LDpred2 non-sparse models derived from Kunkle, Grenier-Boley [38] and sparse models derived from Bellenguez, Küçükali [2], to near-zero (ρ ≈ 0; **Figure 2 panel b**) between Lassosum2 using the Jansen, Savage [39] GWAS and MegaPRS using the Bellenguez, Küçükali [2] GWAS. Moderately positive correlations were observed between LDpred2 models using differing hyperparameters derived from the same Bellenguez, Küçükali [2] GWAS (ρ=0.5; **Figure 2 panel c**), while very strong correlations (ρ=0.9–1; **Figure 2 panels d–e**) were seen among MegaPRS and QuickPRS models derived from the Kunkle, Grenier-Boley [38] GWAS.

To quantify the degree of unique information across the full set of PRS, we applied a greedy selection procedure to identify representative, minimally correlated features across a range of Spearman’s ρ thresholds (**Figure 2 panel f**). At a threshold of ρ<0.3, the PRS set was reduced to ∼15 largely uncorrelated scores per ancestry (range 15-18 across AMR, AFR, EAS, and EUR groups). More permissive correlation thresholds increased the count of distinct PRS, with highly consistent patterns observed across ancestry groups. **Supplementary Table S14** details the number of unique PRS present at each Spearman’s ρ threshold across each genetic ancestry.

### Within-individual PRS variability

Consistent with the wide range in correlation observed between PRS methodologies, individual-level AD risk predictions using those methods also had high variability. Across all 1,752 PRS computed for EUR-ancestry participants (**Supplementary Table S16**; *n*=13,232; 4,392 cases, 8,840 controls), 8,313 individuals (62.8%) reached the 99^th^ percentile according to at least one PRS (range: 1-1,036 PRS in 99^th^ percentile per individual). Among those individuals, 3,078 (37.0%) were AD cases. In contrast, individuals never reaching the 99^th^ percentile (*n*=4,919) were significantly less likely to be an AD case (*n*=1,314; 26.7%) compared to individuals with at least one PRS in the 99^th^ percentile (OR = 1.39; *P* = 4.20 × 10^-34^, chi-square test).

For both EUR and non-EUR ancestries (**Supplementary Tables S17-S19**), extreme within-individual discordance remained common across ancestries; PRS values were reported in both the top and bottom deciles for 96.6% of AMR cases and controls (3,104/3,212), 98.3% of AFR cases and controls (3,335/3,393), 96.7% of EAS cases and controls (2,213/2,289), and 95.8% of EUR cases and controls (12,681/13,232).

**Figure 3** illustrates the extreme range of PRS *z*-scores that can be obtained for the same individual depending on the choice of algorithm and GWAS. For each ancestry, the individual with the maximum PRS range is shown. Of the original 1,752 PRS, we plotted only PRS with ρ < 0.3 to emphasize truly independent signals and to produce a cleaner visualization with minimal overlap. The widest spreads were observed in AFR ancestry (maximum range = 3.303 standard deviations), EUR (2.169 standard deviations), AMR (2.116 standard deviations), and EAS (1.997 standard deviations). **Supplementary Figures S1 and S2** highlight the wide range of risk scores that can occur for an individual, even when the same GWAS summary statistics or algorithm are employed. **Supplementary Figure S3** similarly shows the individual with the minimum *z*-score range for each ancestry (0.392 in AFR, 0.272 in AMR, 0.353 in EAS, and 0.196 in EUR), indicating that even the most stable PRS profiles still exhibit noticeable method-dependent differences in risk.

**Figure 3.**
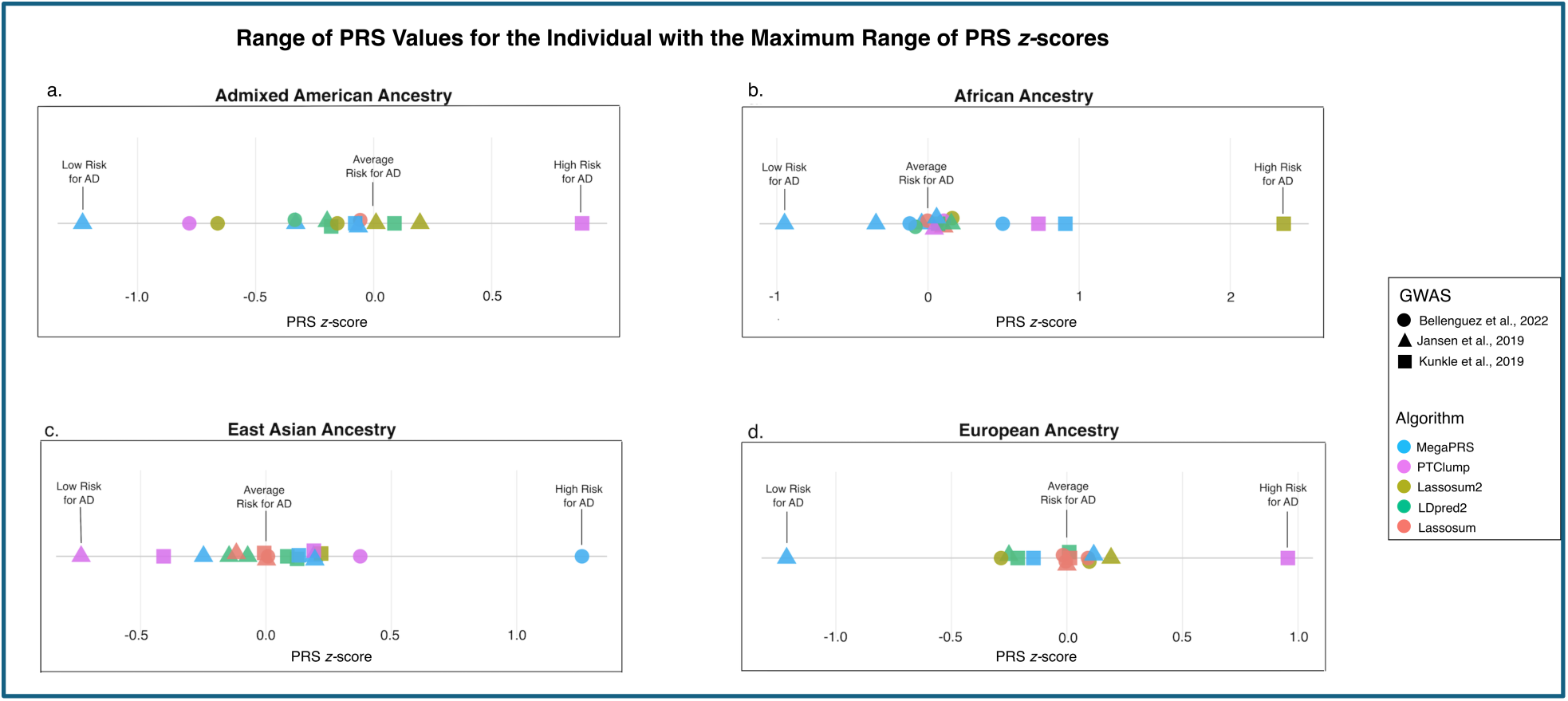
Range of uncorrelated (ρ < 0.3) PRS for AD calculated for the individual with the maximum range of PRS z-scores for each genetic ancestry (A: AMR, B: AFR, C: EAS, D: EUR). PRS z-scores were computed using five different algorithms, indicated by color, and three different GWAS summary statistics, indicated by shape. Substantial variability in PRS values can arise for the same individual depending on the choice of algorithm and GWAS.

### Optimal PRS thresholds and AD risk stratification across ancestries

Since ∼20% of individuals will have dementia by age 85 [55], we decided to optimize ancestry-specific PRS models and thresholds that maximized positive predictive value while ensuring at least 20% of individuals in each ancestry group exceeded the PRS cutoff. This approach allowed us to capture segments of the PRS distribution with meaningfully elevated AD risk while ensuring that the results are broadly applicable, similar to a threshold that might be used in future clinical applications. The optimal PRS constructions varied by ancestry: for AFR, we selected the Bellenguez, Küçükali [2] summary statistics with MegaPRS (PRS #11; LDAK Model 179; threshold PRS ≥ - 0.217); for AMR, PRS #12 using the Kunkle, Grenier-Boley [38] summary statistics with Lassosum (s=1, λ=0.00360; threshold PRS ≥ -0.103); for EAS, PRS #13 using the Jansen, Savage [39] summary statistics with DBSLMM (default heritability scaled by a factor of 1.2; threshold PRS ≥ -0.146); and for EUR, PRS #14 using the Kunkle, Grenier-Boley [38] summary statistics with LDpred2 non-sparse mode (*P* = 0.056, h^2^ = 0.07; threshold PRS ≥ -0.082). We provide full characteristics of these optimal PRS models and thresholds in **Supplementary Table S20**.

We examined the relationship between PRS *z*-scores and AD risk at each PRS decile, stratified by ancestry and *APOE* diplotype, for each of the ancestry-specific models that resulted in the optimal PRS threshold. Lower PRS generally decreased AD risk, while higher PRS generally increased AD risk relative to the *APOE* diplotype for each population (**Figure 4**).

**Figure 4.**
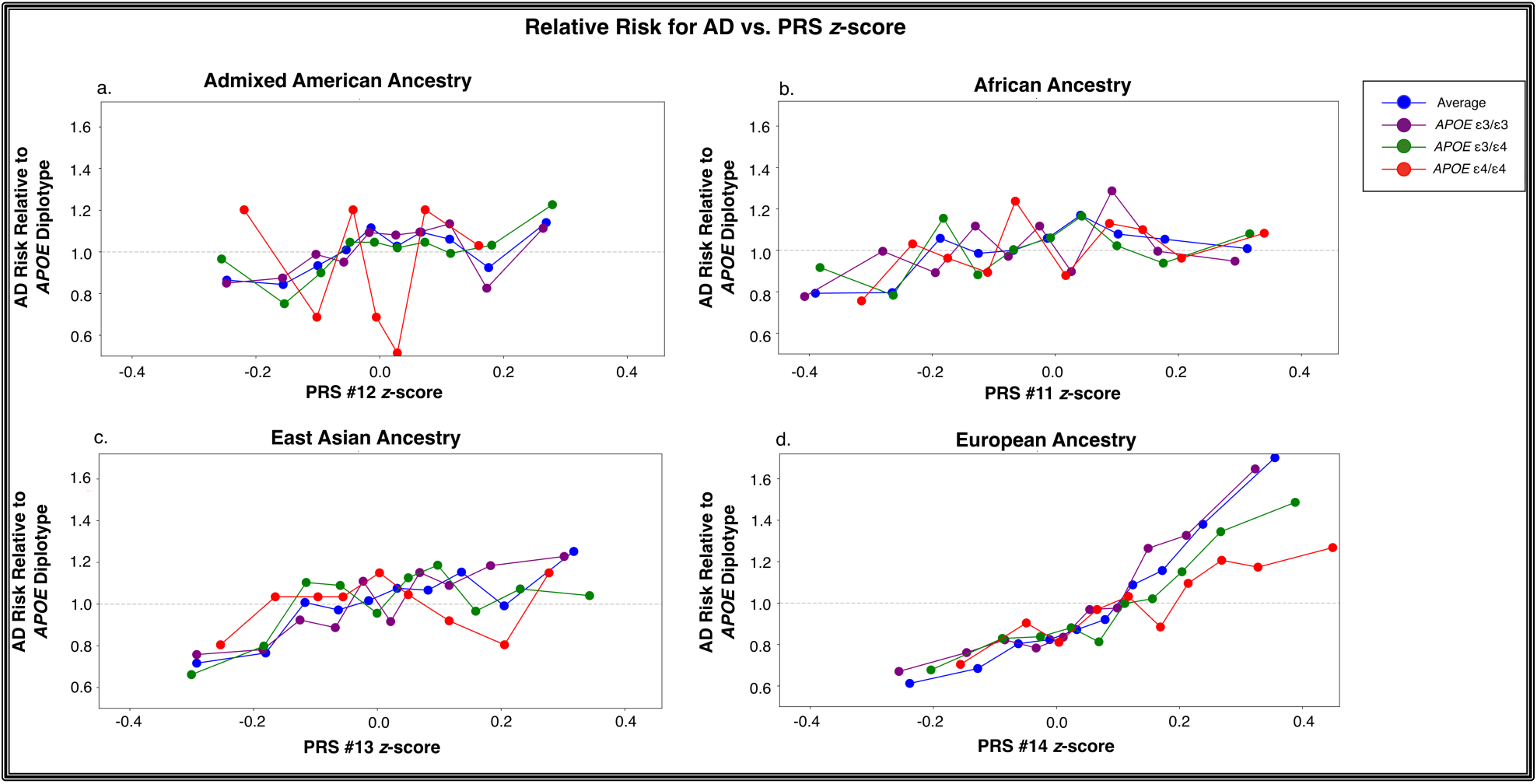
Relative risk for AD for each PRS decile stratified by APOE diplotype and genetic ancestry. Each point shows AD risk for each decile within an APOE diplotype, with each color showing the APOE diplotype. The subplots show the genetic ancestries (a: AMR, b: AFR, c: EAS, d: EUR). The horizontal grey line indicates the average AD risk for each APOE diplotype. Points below that line indicate decreased risk for AD while points above that line indicate increased risk for AD. For each ancestry, the x-axes show standardized PRS values for models labeled PRS #11-#14 (see **Supplementary Table S15** for descriptions of all PRS models). The PRS method producing the highest precision at thresholds including at least 20% of the population was used. Note: Some populations (e.g., AMR) have relatively few individuals with the APOE ε4/ε4 diplotype (n=79 for AMR), resulting in wide variation at some PRS thresholds when stratified by deciles. We opted to include that variation in this plot instead of combining small decile groups to show that AD risk relative to PRS is not always linear and establishing a high versus low risk PRS threshold could resolve issues with potentially overfitting models to a population with limited samples.

Using the optimal thresholds to stratify AD risk for each population, we showed that PRS significantly improve AD risk stratification for each population beyond that of the *APOE* diplotype (AFR: OR = 1.52, *P* = 2.043 × 10^-26^; AMR: OR = 1.47, *P* = 4.565 × 10^-21^; EAS: OR = 1.84, *P* = 6.213 × 10^-40^; EUR: OR = 1.8, *P* = 7.893 × 10^-187^). **Figure 5** illustrates these differences by showing the proportion of AD diagnoses below and above the optimal PRS cutoffs, stratified by ancestry and *APOE* diplotype. AD risk from PRS was largely independent of the *APOE* diplotype, and even increased AD risk in individuals with *APOE* diplotypes carrying the ε2 allele, which is typically considered protective. For example, looking at EUR participants above and below the PRS threshold, AD prevalence rose across all *APOE* diplotypes: ε2/ε2: 0% below the threshold to 11% above; ε2/ε3: ∼0% to 18%; ε2/ε4: 28% to 36%; ε3/ε3: 18% to 28%; ε3/ε4: 32% to 47%; ε4/ε4: 41% to 63%. We observed comparable increases in AD risk above and below each threshold in the AFR, AMR, and EAS populations. Full details of the PRS configurations for each ancestry appear in **Supplementary Table S21**.

**Figure 5.**
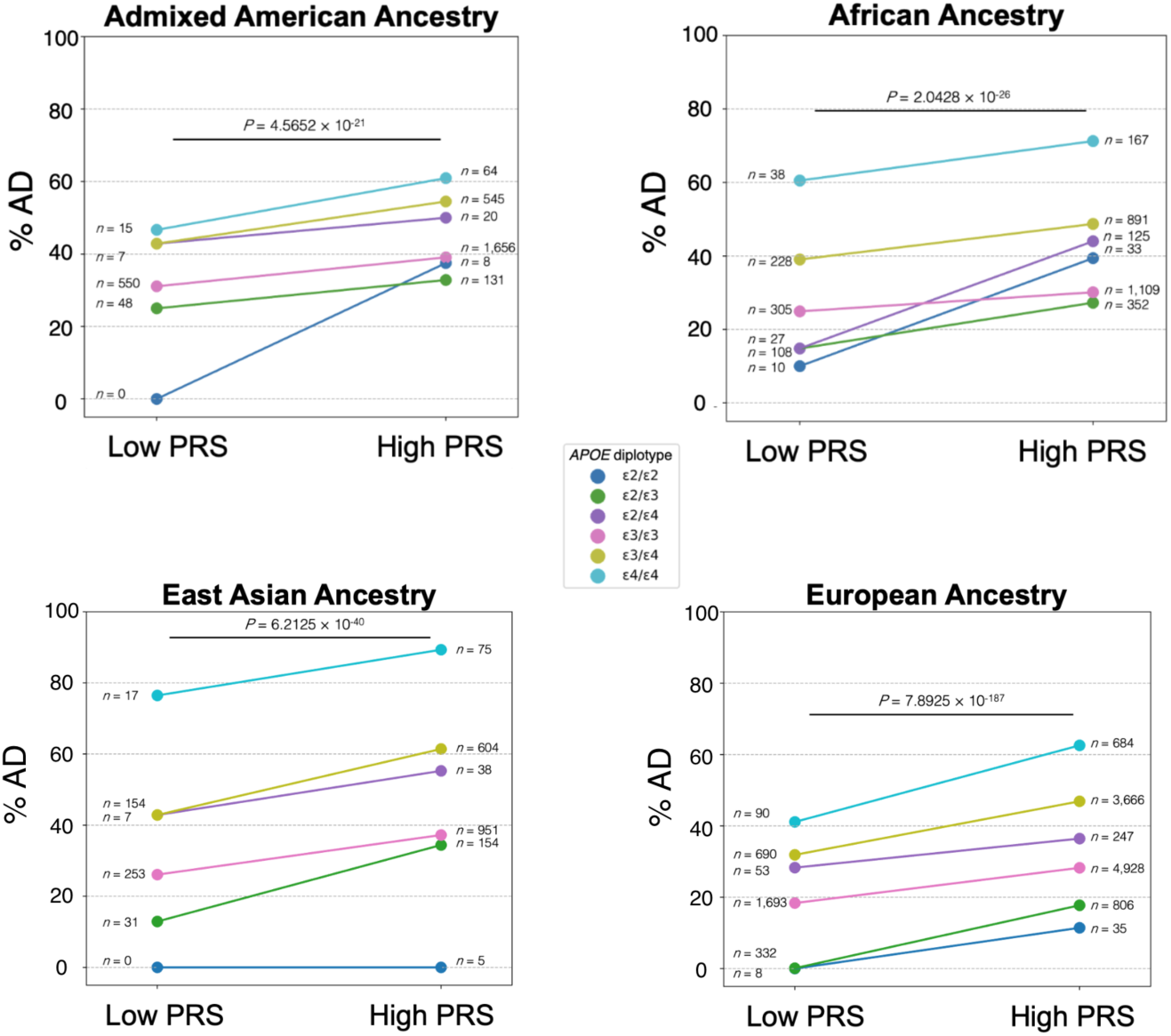
Proportion of AD diagnoses below and above optimal PRS cuto=s, stratified by ancestry and APOE diplotype. Thresholds were selected to maximize precision while including ≥20% of each ancestry group. P-values from chi-square tests indicate highly significant differences in the distribution of AD cases across APOE diplotypes between the low- and high-PRS groups within each ancestry. Descriptions of the PRS configurations (GWAS source, algorithm, and hyperparameters) for each ancestry are included in **Supplementary Table S21**.

### Evaluating the Linearity of PRS-AD Risk Association

In the EUR ancestry, the slope of PRS on AD risk below the threshold of -0.082 was shallow and imprecisely estimated, with a confidence interval spanning the null (OR = 1.18 per unit PRS; 95% CI: 0.44-3.26; *P* = 0.747), while the slope above the threshold was markedly steeper and estimated with substantially greater precision (OR = 15.74 per unit PRS; 95% CI: 11.43-21.73; *P* = 1.858 × 10^-63^). The interaction term, which quantifies how much steeper the PRS slope is above the threshold relative to below it, was highly significant (OR = 13.35; 95% CI: 4.61-37.60; *P* =1.29 × 10^-6^). An LRT confirmed that the interaction model fit the data significantly better than the main-effects model (χ²(1) = 21.85; *P* = 2.96 × 10^-6^), indicating that the PRS-AD relationship is non-linear with a meaningful inflection at the identified threshold. Notably, the below-threshold estimate carried substantially greater uncertainty than the above-threshold estimate, consistent with differences in sample size and case representation between the two segments. This pattern is illustrated in **Figure 6**, where a LOESS smoother fit to the full data without imposing any threshold shows a natural inflection near -0.082, corroborating the threshold as a data-driven boundary rather than an arbitrary cutoff. As expected, the *APOE* diplotype was a significant independent covariate of the logistic regression, with the ε4 allele increasing AD risk (ε4/ε4: OR = 3.14; 95% CI: 2.67-3.68; ε2/ε3: OR = 0.55, 95% CI: 0.46-0.65). The interaction was not significant in any of the three non-European ancestry groups (**Supplementary Table S22**), with slopes below and above the threshold remaining comparable within each group.

**Figure 6.**
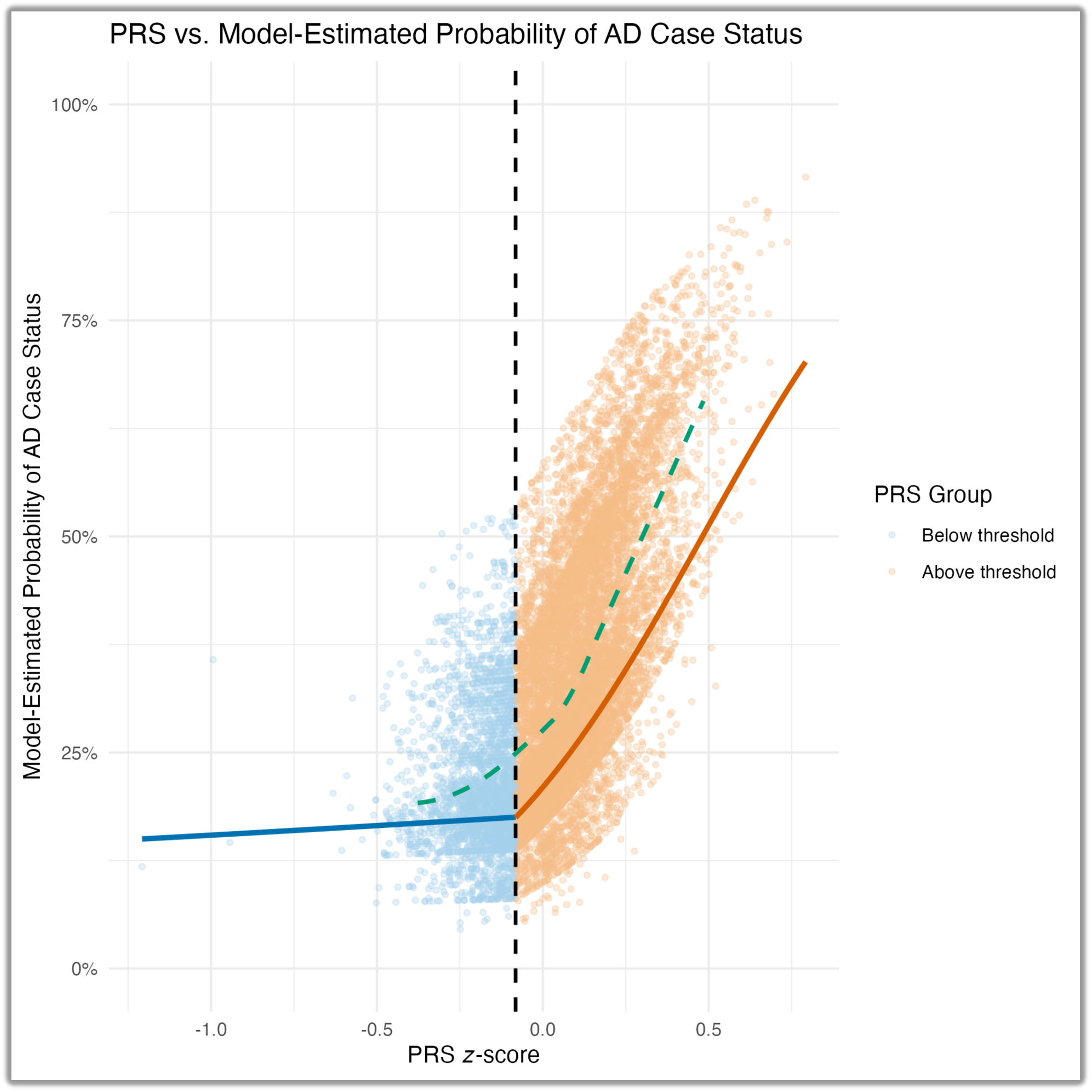
Model-estimated probability of Alzheimer’s disease case status as a function of PRS z-score in individuals of EUR ancestry. Each point represents one individual, colored by PRS group (blue: below threshold, PRS < -0.082; orange: above threshold, PRS ≥ -0.082). The vertical dashed line indicates the identified threshold of - 0.082. Predicted probabilities reflect the logistic regression interaction model’s estimated likelihood that an individual is an AD case given their PRS, age, sex, and APOE diplotype; they do not represent absolute population-level AD risk, and vertical dispersion of points at a given PRS value reflects variation in these covariates— particularly APOE diplotype—across individuals. Solid lines represent model-derived predicted probabilities fit separately below (blue) and above (orange) the threshold, with age, sex, and APOE diplotype held at reference (median age, female sex, ε3/ε3 diplotype) to isolate the contribution of PRS. The dashed green line is a LOESS smoother fit to the full data across the 1^st^-99^th^ percentile of PRS without imposing any threshold, shown as a data-driven reference. Its natural inflection near -0.082 corroborates the threshold as a meaningful boundary rather than an arbitrary cutoff.

### Evaluating the effects of GWAS sample overlap

We evaluated the impact of potential sample overlap between the ADSP R5 WGS cohort and the GWAS summary statistics using two complementary analyses. We based all primary PRS performance metrics, benchmarking comparisons, and identification of the top-performing models on the full ADSP sample, as excluding likely-overlapped individuals did not materially change performance rankings or overall conclusions.

For the 1,168 PRS derived from Kunkle, Grenier-Boley [38] and Jansen, Savage [39] summary statistics, we compared PRS distributions between the likely-overlapped subgroup (participants with prior WES) and the unlikely-overlapped subgroup (WGS-only participants). We took a conservative approach by evaluating changes in the median and distributions using both the Mann-Whitney U and Kolmogorov-Smirnov (KS) tests. For the EUR population, 248 of 1,168 PRS showed significant distributional shifts with the Mann-Whitney U test and 262 by the KS test (*P* < 4.281 x 10^-5^ after Bonferroni correction for 1,168 tests), with maximum KS statistics ranging from 0.085 to 0.15— indicating low absolute differences despite large sample sizes. In AMR individuals, 181 PRS had significantly different distributions in the cohorts likely used in the GWAS compared to the other WGS samples with Mann-Whitney U and 173 by KS test (Bonferroni-corrected), but with notably larger maximum KS statistics (0.25-0.53). No PRS reached Bonferroni-corrected significance in AFR individuals for either test.

For the 584 PRS derived from Bellenguez, Küçükali [2] summary statistics, we compared the likely-overlapped subgroup (ADGC cohorts) versus the unlikely-overlapped subgroup (non-ADGC individuals). In the EUR population, 449 of 584 PRS had significantly different PRS distributions using the Mann-Whitney U test and 389 had significantly different distributions by KS test (*P* < 8.562 x 10^-5^ after Bonferroni correction for 584 tests). However, these shifts were small in absolute magnitude, with maximum KS statistics ranging from 0.042 to 0.066, indicating very small distributional differences despite the high proportion of significant tests and large sample sizes. In the AMR population, fewer PRS showed significance (43 by Mann-Whitney U and 23 by KS test after correction), but the shifts were larger in magnitude, with maximum KS statistics ranging from 0.25 to 0.51. No PRS reached Bonferroni-corrected significance in AFR individuals for ADGC stratification (**Supplementary Tables S5-S12**).

The PRS method ultimately selected for the primary analyses in AFR and AMR ancestries did not yield statistically significant p-values in either the KS or Mann-Whitney U tests, indicating the samples likely used in the original GWAS were not significantly different than the ADSP samples used in validation. In contrast, the EUR-ancestry PRS had significant p-values, although the KS statistic was low (0.12), indicating that although there were significant differences in the sample distributions between the EUR GWAS samples and the rest of the EUR samples in the ADSP, the magnitude of those differences was low. Importantly, the top-performing PRS configuration remained unchanged between the “unlikely overlapped” ADSP subsample and all EUR individuals in the ADSP, supporting the robustness of our primary choice regardless of whether GWAS-overlapping samples are included in the PRS benchmarks. Overlap exploration was not performed for the EAS group due to the extremely limited availability of overlapping data (ADGC: 37/2,289; WES: 1/2,289).

## Discussion

In this study, we conducted an extensive evaluation of AD PRS across GWAS sources, algorithms, and hyperparameter configurations using ADSP R5 dataset. By systematically comparing 1,752 PRS models across four genetically inferred ancestries, we demonstrate that methodological choices profoundly influence PRS behavior and performance. We further show that when optimally parameterized and evaluated using threshold-based approaches, PRS significantly improve AD risk stratification beyond *APOE* diplotyping across AMR, AFR, EAS, and EUR populations, even when the base GWAS summary statistics were based on an EUR cohort.

A central finding of this study is the remarkable range of observed PRS depending on methodological choice, even after standardizing to a reference population. Despite all scores nominally representing AD PRS, correlations among scores ranged from strongly positive to moderately negative for the same individuals. Those differences arose not only across GWAS summary statistics (as seen in previous studies [30, 56]), but also across algorithms and hyperparameter choices applied to the same GWAS. Our findings show that PRS constructed using different algorithms and summary statistics can result in AD risk predictions that are either perfectly correlated, uncorrelated, or even negatively correlated for the same individuals, underscoring that conclusions derived from PRS conducted using different methodologies are not interchangeable and simply specifying the GWAS and PRS method used to calculate the AD PRS is not sufficient without also including the hyperparameters used for score calculation.

Divergent findings across AD PRS studies may reflect methodological choice rather than true biological inconsistency. In this context, negative or null results should not necessarily be interpreted as evidence against PRS utility, without first evaluating the potential consequences of suboptimal algorithm selection, insufficient hyperparameter tuning, or mismatched evaluation frameworks. Our correlation-based analyses demonstrate that many commonly used PRS capture overlapping or redundant information, while others reflect largely independent genetic components of AD susceptibility, emphasizing the need for careful method selection rather than arbitrary choices.

Current PRS benchmarking practices, including those adopted by the PGS Catalog [57], primarily rely on linear metrics such as effect size per standard deviation, incremental R^2^, or area under the receiver operating characteristic curve (AUROC) [30, 58]. These metrics implicitly assume that genetic risk increases approximately linearly across the PRS distribution. However, **Figure 6** challenges this assumption for late-onset AD in individuals of EUR ancestry, showing that the slope of the PRS-AD association differs significantly above versus below the optimal threshold (interaction OR = 13.35, 95% CI: 4.61-37.60, *P* = 1.29 × 10^-6^). These findings indicate that in EUR samples, AD PRS do not follow a linear model, but instead exhibit either nonlinear or threshold-dependent effects in which PRS becomes markedly more informative above a critical score value.

However, the below-threshold slope itself was estimated with considerably less precision than the above-threshold slope, with a confidence interval spanning the null. This asymmetry is consistent with the substantially smaller number of AD cases available to inform the below-threshold estimate. Of the 13,232 individuals in the EUR ancestry cohort, only 2,880 (21.8%) fell below the PRS threshold, including just 620 AD cases, compared to 3,772 cases among the 10,352 individuals above the threshold. Because the precision of a logistic regression coefficient depends on the number of observed events rather than sample size alone, the below-threshold estimate carries correspondingly greater sampling variance. This pattern likely reflects reduced statistical power within the below-threshold segment rather than evidence against a true PRS effect in that range, and the estimate should be interpreted with appropriate caution. Larger below-threshold samples would help clarify whether the apparent flattening of the PRS-AD relationship below the threshold reflects a genuine plateau in risk or is instead an artifact of limited precision in this segment.

Despite the reduced precision of the below-threshold estimate, the overall pattern of a significantly steeper PRS-AD slope above the threshold is consistent with a liability-threshold model of disease risk [59], in which genetic burden accumulates until a critical point is reached, after which disease susceptibility increases sharply. While such models are well established in classical quantitative genetics, they have received comparatively little attention in contemporary PRS evaluation frameworks. As a result, reliance on linear metrics may systematically underestimate the clinical and translational relevance of PRS for disorders such as AD, where genetic risk appears concentrated within the extreme tail rather than distributed evenly across the population.

Notably, the significant non-linearity detected in the EUR group was not found in the AMR, AFR, or EAS groups (**Supplementary Table S22**), where slopes below and above the optimal threshold remained comparable. This difference between EUR and other populations may reflect the attenuated PRS signal in non-EUR samples resulting from the predominantly EUR composition of the underlying GWAS, which reduces statistical power to detect slope differences, particularly given the smaller sample sizes in non-EUR groups. Threshold-based stratification nonetheless successfully stratified individuals with elevated AD prevalence across all four ancestry groups (**Figures 4 and 5**), suggesting that the threshold-based approach can appropriately stratify risk groups even if the slope of PRS risk significantly changes only in the EUR population.

Thresholds selected in this study were not restricted to ultra-rare extremes but instead were chosen to include at least 20% of individuals within each ancestry. Even within those relatively broad strata, PRS identified groups with markedly higher AD prevalence across nearly all *APOE* diplotypes, demonstrating that PRS capture genetic information that is largely independent of the *APOE* diplotype.

These results help reconcile long-standing debates regarding the incremental value of PRS beyond *APOE.* When evaluated linearly across all possible PRS thresholds, PRS may appear to add modest explanatory power beyond that of *APOE* [30, 60] (**Figure 4**). However, our evaluations show that even when PRS may appear to have limited utility (e.g., in the AMR and AFR populations), a threshold-based framework stratified by *APOE* (**Figure 5)** may be more appropriate to predict AD cases and controls. This distinction is critical for interpreting prior studies and for defining appropriate expectations for PRS utility in AD.

The most immediate implication of these findings lies in cohort risk stratification rather than individual-level diagnosis. In the context of AD prevention and disease-modifying trials, efficient identification of high-risk individuals remains a major challenge [61]. Threshold-based PRS offer a scalable approach for identifying individuals who might benefit from additional cognitive screening and enriching for at-risk individuals in pre-symptomatic clinical trials.

Importantly, our results demonstrate that such enrichment is achievable across multiple ancestry groups, even when PRS are derived from primarily EUR-based GWAS. Although predictive performance remains attenuated in non-EUR populations, and formal non-linearity testing was limited by GWAS transferability and sample size constraints, appropriately tuned PRS nonetheless identify high-risk strata with significantly elevated AD prevalence. These findings argue against the notion that PRS are categorically uninformative outside EUR ancestry and instead support continued methodological refinement alongside ongoing efforts to diversify GWAS representation.

We acknowledge that partial sample overlap existed between the ADSP dataset and the GWAS summary statistics used for PRS construction, which resulted in statistically significant differences in PRS distributions between likely overlapped and non-overlapped individuals. In EUR ancestry participants, these differences were generally small in magnitude, and the relative ranking and performance of top-performing PRS models remained stable after exclusion of likely overlapped samples, indicating that GWAS sample overlap did not materially influence the primary conclusions of this study.

In contrast, larger distributional shifts were observed in AMR participants, despite fewer comparisons reaching statistical significance. This pattern may reflect increased sensitivity to overlap in admixed populations, where differences in allele frequencies and LD structure can amplify subtle correlations between discovery and target datasets, especially when the sample size of the AMR population is smaller than that of the EUR population. In addition, overlap effects were more pronounced for PRS derived from the Bellenguez, Küçükali [2] GWAS, which incorporated a large number of ADGC cohorts that also contributed samples to the ADSP. Compared with earlier GWAS that included more limited WES overlap, the greater scale of shared cohorts and closer alignment of diagnostic and sequencing frameworks likely increased detectable overlap effects. Importantly, exclusion of likely overlapped individuals did not alter overall PRS performance patterns or model rankings, supporting the robustness of the study’s central findings.

Our analyses further highlight that reported race does not reliably correspond to genetically inferred ancestry within the ADSP dataset. Concordance varied substantially across groups, and only ∼65% of individuals within a reported race category shared the most common genetically inferred ancestry for that category. These discrepancies have direct implications for PRS analyses, as ancestry mismatches can introduce bias through differences in allele frequencies and LD structure [62]. Accordingly, PRS analyses should rely on genetically inferred ancestry rather than self-reported race. PRS that do not stratify by genetic ancestry risk conflating social and genetic constructs, potentially obscuring true genetic effects and reducing reproducibility across studies.

We recognize that although extensive hyperparameter tuning was performed, PRS optimization remains constrained by the available GWAS summary statistics, which are still disproportionately derived from EUR populations. it is clear that current PRS methods, while better than *APOE* baselines, still fall short of modeling the 60-80% of AD risk that is thought to be heritable [1].

We opted for a more inclusive assessment of PRS algorithmic comparisons by looking at only common variant associations with AD in a broad cohort that included both clinical and autopsy-confirmed AD cases. Including rare variants, epistatic interactions, and gene-environment interactions in the risk model is likely to improve performance metrics.

It is also important to note that only ∼40% of AD-type dementia is attributable to AD neuropathologic changes [63], so clinical AD cases likely have different genetic risk factors than autopsy-confirmed AD cases and could impact PRS accuracy. Additionally, lifetime cumulative risk models for dementia show that dementia risk increases from ∼4% by age 75 to ∼20% by age 85, and ∼40% by age 95 [55], indicating that many of the younger controls in the ADSP cohort might develop AD later in life. Refining the AD phenotype by including only autopsy-confirmed cases and further stratifying analyses by age are likely to improve risk prediction. Future work should also prioritize the development of ancestry-diverse GWAS, improved LD reference panels, and developing standardized frameworks for threshold-based PRS evaluation.

In summary, our results demonstrate that polygenic risk for AD is both substantial and informative for cohort-level risk stratification, but highly sensitive to methodological choices and evaluation frameworks. Cross-study PRS should not be treated as interchangeable unless they use the same GWAS, PRS algorithm, and hyperparameters. Additionally, PRS evaluated solely through linear metrics may underestimate PRS importance for risk stratification. These PRS benchmarks show which algorithmic configurations are best at predicting AD case status in the ADSP, provide guidance on how to interpret the underlying genetic architecture of AD encapsulated in PRS, and reveal meaningful risk stratification beyond *APOE* diplotyping across multiple ancestries.

## Data Availability

Access to the ADSP is controlled by The National Institute on Aging Genetics of Alzheimer's Disease (NIAGADS). All scripts used to analyze the data are freely available at https://github.com/jmillerlab/prs_comparisons.

## Acknowledgements

The Alzheimer’s Disease Sequencing Project (ADSP) is comprised of two Alzheimer’s Disease (AD) genetics consortia and three National Human Genome Research Institute (NHGRI) funded Large Scale Sequencing and Analysis Centers (LSAC). The two AD genetics consortia are the Alzheimer’s Disease Genetics Consortium (ADGC) funded by NIA (U01 AG032984), and the Cohorts for Heart and Aging Research in Genomic Epidemiology (CHARGE) funded by NIA (R01 AG033193), the National Heart, Lung, and Blood Institute (NHLBI), other National Institute of Health (NIH) institutes and other foreign governmental and non-governmental organizations. The Discovery Phase analysis of sequence data is supported through UF1AG047133 (to Drs. Schellenberg, Farrer, Pericak-Vance, Mayeux, and Haines); U01AG049505 to Dr. Seshadri; U01AG049506 to Dr. Boerwinkle; U01AG049507 to Dr. Wijsman; and U01AG049508 to Dr. Goate and the Discovery Extension Phase analysis is supported through U01AG052411 to Dr. Goate, U01AG052410 to Dr. Pericak-Vance and U01 AG052409 to Drs. Seshadri and Fornage.

Sequencing for the Follow Up Study (FUS) is supported through U01AG057659 (to Drs. PericakVance, Mayeux, and Vardarajan) and U01AG062943 (to Drs. Pericak-Vance and Mayeux). Data generation and harmonization in the Follow-up Phase is supported by U54AG052427 (to Drs. Schellenberg and Wang). The FUS Phase analysis of sequence data is supported through U01AG058589 (to Drs. Destefano, Boerwinkle, De Jager, Fornage, Seshadri, and Wijsman), U01AG058654 (to Drs. Haines, Bush, Farrer, Martin, and Pericak-Vance), U01AG058635 (to Dr. Goate), RF1AG058066 (to Drs. Haines, Pericak-Vance, and Scott), RF1AG057519 (to Drs. Farrer and Jun), R01AG048927 (to Dr. Farrer), and RF1AG054074 (to Drs. Pericak-Vance and Beecham).

The ADGC cohorts include: Adult Changes in Thought (ACT) (U01 AG006781, U19 AG066567), the Alzheimer’s Disease Research Centers (ADRC) (P30 AG062429, P30 AG066468, P30 AG062421, P30 AG066509, P30 AG066514, P30 AG066530, P30 AG066507, P30 AG066444, P30 AG066518, P30 AG066512, P30 AG066462, P30 AG072979, P30 AG072972, P30 AG072976, P30 AG072975, P30 AG072978, P30 AG072977, P30 AG066519, P30 AG062677, P30 AG079280, P30 AG062422, P30 AG066511, P30 AG072946, P30 AG062715, P30 AG072973, P30 AG066506, P30 AG066508, P30 AG066515, P30 AG072947, P30 AG072931, P30 AG066546, P20 AG068024, P20 AG068053, P20 AG068077, P20 AG068082, P30 AG072958, P30 AG072959), the Chicago Health and Aging Project (CHAP) (R01 AG11101, RC4 AG039085, K23 AG030944), Indiana Memory and Aging Study (IMAS) (R01 AG019771), Indianapolis Ibadan (R01 AG009956, P30 AG010133), the Memory and Aging Project (MAP) (R01 AG17917), Mayo Clinic (MAYO) (R01 AG032990, U01 AG046139, R01 NS080820, RF1 AG051504, P50 AG016574), Mayo Parkinson’s Disease controls (NS039764, NS071674, 5RC2HG005605), University of Miami (R01 AG027944, R01 AG028786, R01 AG019085, IIRG09133827, A2011048), the Multi-Institutional Research in Alzheimer’s Genetic Epidemiology Study (MIRAGE) (R01 AG09029, R01 AG025259), the National Centralized Repository for Alzheimer’s Disease and Related Dementias (NCRAD) (U24 AG021886), the National Institute on Aging Late Onset Alzheimer’s Disease Family Study (NIA-LOAD) (U24 AG056270), the Religious Orders Study (ROS) (P30 AG10161, R01 AG15819), the Texas Alzheimer’s Research and Care Consortium (TARCC) (funded by the Darrell K Royal Texas Alzheimer’s Initiative), Vanderbilt University/Case Western Reserve University (VAN/CWRU) (R01 AG019757, R01 AG021547, R01 AG027944, R01 AG028786, P01 NS026630, and Alzheimer’s Association), the Washington Heights-Inwood Columbia Aging Project (WHICAP) (RF1 AG054023), the University of Washington Families (VA Research Merit Grant, NIA: P50AG005136, R01AG041797, NINDS: R01NS069719), the Columbia University Hispanic Estudio Familiar de Influencia Genetica de Alzheimer (EFIGA) (RF1 AG015473), the University of Toronto (UT) (funded by Wellcome Trust, Medical Research Council, Canadian Institutes of Health Research), and Genetic Differences (GD) (R01 AG007584). The CHARGE cohorts are supported in part by National Heart, Lung, and Blood Institute (NHLBI) infrastructure grant HL105756 (Psaty), RC2HL102419 (Boerwinkle) and the neurology working group is supported by the National Institute on Aging (NIA) R01 grant AG033193.

The CHARGE cohorts participating in the ADSP include the following: Austrian Stroke Prevention Study (ASPS), ASPS-Family study, and the Prospective Dementia Registry-Austria (ASPS/PRODEM-Aus), the Atherosclerosis Risk in Communities (ARIC) Study, the Cardiovascular Health Study (CHS), the Erasmus Rucphen Family Study (ERF), the Framingham Heart Study (FHS), and the Rotterdam Study (RS). ASPS is funded by the Austrian Science Fond (FWF) grant number P20545-P05 and P13180 and the Medical University of Graz. The ASPS-Fam is funded by the Austrian Science Fund (FWF) project I904), the EU Joint Programme – Neurodegenerative Disease Research (JPND) in frame of the BRIDGET project (Austria, Ministry of Science) and the Medical University of Graz and the Steiermärkische Krankenanstalten Gesellschaft. PRODEM-Austria is supported by the Austrian Research Promotion agency (FFG) (Project No. 827462) and by the Austrian National Bank (Anniversary Fund, project 15435. ARIC research is carried out as a collaborative study supported by NHLBI contracts (HHSN268201100005C, HHSN268201100006C, HHSN268201100007C, HHSN268201100008C, HHSN268201100009C, HHSN268201100010C, HHSN268201100011C, and HHSN268201100012C). Neurocognitive data in ARIC is collected by U01 2U01HL096812, 2U01HL096814, 2U01HL096899, 2U01HL096902, 2U01HL096917 from the NIH (NHLBI, NINDS, NIA and NIDCD), and with previous brain MRI examinations funded by R01-HL70825 from the NHLBI. CHS research was supported by contracts HHSN268201200036C, HHSN268200800007C, N01HC55222, N01HC85079, N01HC85080, N01HC85081, N01HC85082, N01HC85083, N01HC85086, and grants U01HL080295 and U01HL130114 from the NHLBI with additional contribution from the National Institute of Neurological Disorders and Stroke (NINDS). Additional support was provided by R01AG023629, R01AG15928, and R01AG20098 from the NIA. FHS research is supported by NHLBI contracts N01-HC-25195 and HHSN268201500001I. This study was also supported by additional grants from the NIA (R01s AG054076, AG049607 and AG033040 and NINDS (R01 NS017950). The ERF study as a part of EUROSPAN (European Special Populations Research Network) was supported by European Commission FP6 STRP grant number 018947 (LSHG-CT-2006-01947) and also received funding from the European Community’s Seventh Framework Programme (FP7/2007-2013)/grant agreement HEALTH-F4-2007-201413 by the European Commission under the programme “Quality of Life and Management of the Living Resources” of 5th Framework Programme (no. QLG2-CT-2002-01254). High-throughput analysis of the ERF data was supported by a joint grant from the Netherlands Organization for Scientific Research and the Russian Foundation for Basic Research (NWO-RFBR 047.017.043). The Rotterdam Study is funded by Erasmus Medical Center and Erasmus University, Rotterdam, the Netherlands Organization for Health Research and Development (ZonMw), the Research Institute for Diseases in the Elderly (RIDE), the Ministry of Education, Culture and Science, the Ministry for Health, Welfare and Sports, the European Commission (DG XII), and the municipality of Rotterdam. Genetic data sets are also supported by the Netherlands Organization of Scientific Research NWO Investments (175.010.2005.011, 911-03-012), the Genetic Laboratory of the Department of Internal Medicine, Erasmus MC, the Research Institute for Diseases in the Elderly (014-93-015; RIDE2), and the Netherlands Genomics Initiative (NGI)/Netherlands Organization for Scientific Research (NWO) Netherlands Consortium for Healthy Aging (NCHA), project 050-060-810. All studies are grateful to their participants, faculty and staff. The content of these manuscripts is solely the responsibility of the authors and does not necessarily represent the official views of the National Institutes of Health or the U.S. Department of Health and Human Services.

The FUS cohorts include: the Alzheimer’s Disease Research Centers (ADRC) (P30 AG062429, P30 AG066468, P30 AG062421, P30 AG066509, P30 AG066514, P30 AG066530, P30 AG066507, P30 AG066444, P30 AG066518, P30 AG066512, P30 AG066462, P30 AG072979, P30 AG072972, P30 AG072976, P30 AG072975, P30 AG072978, P30 AG072977, P30 AG066519, P30 AG062677, P30 AG079280, P30 AG062422, P30 AG066511, P30 AG072946, P30 AG062715, P30 AG072973, P30 AG066506, P30 AG066508, P30 AG066515, P30 AG072947, P30 AG072931, P30 AG066546, P20 AG068024, P20 AG068053, P20 AG068077, P20 AG068082, P30 AG072958, P30 AG072959), Alzheimer’s Disease Neuroimaging Initiative (ADNI) (U19AG024904), Amish Protective Variant Study (RF1AG058066), Cache County Study (R01AG11380, R01AG031272, R01AG21136, RF1AG054052), Case Western Reserve University Brain Bank (CWRUBB) (P50AG008012), Case Western Reserve University Rapid Decline (CWRURD) (RF1AG058267, NU38CK000480), CubanAmerican Alzheimer’s Disease Initiative (CuAADI) (3U01AG052410), Estudio Familiar de Influencia Genetica en Alzheimer (EFIGA) (5R37AG015473, RF1AG015473, R56AG051876), Genetic and Environmental Risk Factors for Alzheimer Disease Among African Americans Study (GenerAAtions) (2R01AG09029, R01AG025259, 2R01AG048927), Gwangju Alzheimer and Related Dementias Study (GARD) (U01AG062602), Hillblom Aging Network (2014-A-004-NET, R01AG032289, R01AG048234), Hussman Institute for Human Genomics Brain Bank (HIHGBB) (R01AG027944, Alzheimer’s Association “Identification of Rare Variants in Alzheimer Disease”), Ibadan Study of Aging (IBADAN) (5R01AG009956), Longevity Genes Project (LGP) and LonGenity (R01AG042188, R01AG044829, R01AG046949, R01AG057909, R01AG061155, P30AG038072), Mexican Health and Aging Study (MHAS) (R01AG018016), Multi-Institutional Research in Alzheimer’s Genetic Epidemiology (MIRAGE) (2R01AG09029, R01AG025259, 2R01AG048927), Northern Manhattan Study (NOMAS) (R01NS29993), Peru Alzheimer’s Disease Initiative (PeADI) (RF1AG054074), Puerto Rican 1066 (PR1066) (Wellcome Trust (GR066133/GR080002), European Research Council (340755)), Puerto Rican Alzheimer Disease Initiative (PRADI) (RF1AG054074), Reasons for Geographic and Racial Differences in Stroke (REGARDS) (U01NS041588), Research in African American Alzheimer Disease Initiative (REAAADI) (U01AG052410), the Religious Orders Study (ROS) (P30 AG10161, P30 AG72975, R01 AG15819, R01 AG42210), the RUSH Memory and Aging Project (MAP) (R01 AG017917, R01 AG42210, Stanford Extreme Phenotypes in AD (R01AG060747), University of Miami Brain Endowment Bank (MBB), University of Miami/Case Western/North Carolina A&T African American (UM/CASE/NCAT) (U01AG052410, R01AG028786), Wisconsin Registry for Alzheimer’s Prevention (WRAP) (R01AG027161 and R01AG054047), Mexico-Southern California Autosomal Dominant Alzheimer’s Disease Consortium (R01AG069013), Center for Cognitive Neuroscience and Aging (R01AG047649), and the A4 Study (R01AG063689, U19AG010483 and U24AG057437).

The four LSACs are: the Human Genome Sequencing Center at the Baylor College of Medicine (U54 HG003273), the Broad Institute Genome Center (U54HG003067), The American Genome Center at the Uniformed Services University of the Health Sciences (U01AG057659), and the Washington University Genome Institute (U54HG003079). Genotyping and sequencing for the ADSP FUS is also conducted at John P. Hussman Institute for Human Genomics (HIHG) Center for Genome Technology (CGT).

Biological samples and associated phenotypic data used in primary data analyses were stored at Study Investigators institutions, and at the National Centralized Repository for Alzheimer’s Disease and Related Dementias (NCRAD, U24AG021886) at Indiana University funded by NIA. Associated Phenotypic Data used in primary and secondary data analyses were provided by Study Investigators, the NIA funded Alzheimer’s Disease Centers (ADCs), and the National Alzheimer’s Coordinating Center (NACC, U24AG072122) and the National Institute on Aging Genetics of Alzheimer’s Disease Data Storage Site (NIAGADS, U24AG041689) at the University of Pennsylvania, funded by NIA. Harmonized phenotypes were provided by the ADSP Phenotype Harmonization Consortium (ADSP-PHC), funded by NIA (U24 AG074855, U01 AG068057 and R01 AG059716) and Ultrascale Machine Learning to Empower Discovery in Alzheimer’s Disease Biobanks (AI4AD, U01 AG068057). This research was supported in part by the Intramural Research Program of the National Institutes of health, National Library of Medicine. Contributors to the Genetic Analysis Data included Study Investigators on projects that were individually funded by NIA, and other NIH institutes, and by private U.S. organizations, or foreign governmental or nongovernmental organizations.

The ADSP Phenotype Harmonization Consortium (ADSP-PHC) is funded by NIA (U24 AG074855, U01 AG068057 and R01 AG059716). The harmonized cohorts within the ADSP-PHC include: the Anti-Amyloid Treatment in Asymptomatic Alzheimer’s study (A4 Study), a secondary prevention trial in preclinical Alzheimer’s disease, aiming to slow cognitive decline associated with brain amyloid accumulation in clinically normal older individuals. The A4 Study is funded by a public-private-philanthropic partnership, including funding from the National Institutes of Health-National Institute on Aging, Eli Lilly and Company, Alzheimer’s Association, Accelerating Medicines Partnership, GHR Foundation, an anonymous foundation and additional private donors, with in-kind support from Avid and Cogstate. The companion observational Longitudinal Evaluation of Amyloid Risk and Neurodegeneration (LEARN) Study is funded by the Alzheimer’s Association and GHR Foundation. The A4 and LEARN Studies are led by Dr. Reisa Sperling at Brigham and Women’s Hospital, Harvard Medical School and Dr. Paul Aisen at the Alzheimer’s Therapeutic Research Institute (ATRI), University of Southern California. The A4 and LEARN Studies are coordinated by ATRI at the University of Southern California, and the data are made available through the Laboratory for Neuro Imaging at the University of Southern California. The participants screening for the A4 Study provided permission to share their de-identified data in order to advance the quest to find a successful treatment for Alzheimer’s disease. We would like to acknowledge the dedication of all the participants, the site personnel, and all of the partnership team members who continue to make the A4 and LEARN Studies possible. The complete A4 Study Team list is available on: a4study.org/a4-study-team.; the Adult Changes in Thought study (ACT), U01 AG006781, U19 AG066567; Alzheimer’s Disease Neuroimaging Initiative (ADNI): Data collection and sharing for this project was funded by the Alzheimer’s Disease Neuroimaging Initiative (ADNI) (National Institutes of Health Grant U01 AG024904) and DOD ADNI (Department of Defense award number W81XWH-12-2-0012). ADNI is funded by the National Institute on Aging, the National Institute of Biomedical Imaging and Bioengineering, and through generous contributions from the following: AbbVie, Alzheimer’s Association; Alzheimer’s Drug Discovery Foundation; Araclon Biotech; BioClinica, Inc.; Biogen; Bristol-Myers Squibb Company; CereSpir, Inc.; Cogstate; Eisai Inc.; Elan Pharmaceuticals, Inc.; Eli Lilly and Company; EuroImmun; F. Hoffmann-La Roche Ltd and its affiliated company Genentech, Inc.; Fujirebio; GE Healthcare; IXICO Ltd.;Janssen Alzheimer Immunotherapy Research & Development, LLC.; Johnson & Johnson Pharmaceutical Research & Development LLC.; Lumosity; Lundbeck; Merck & Co., Inc.;Meso Scale Diagnostics, LLC.; NeuroRx Research; Neurotrack Technologies; Novartis Pharmaceuticals Corporation; Pfizer Inc.; Piramal Imaging; Servier; Takeda Pharmaceutical Company; and Transition Therapeutics. The Canadian Institutes of Health Research is providing funds to support ADNI clinical sites in Canada. Private sector contributions are facilitated by the Foundation for the National Institutes of Health (www.fnih.org). The grantee organization is the Northern California Institute for Research and Education, and the study is coordinated by the Alzheimer’s Therapeutic Research Institute at the University of Southern California. ADNI data are disseminated by the Laboratory for Neuro Imaging at the University of Southern California; Estudio Familiar de Influencia Genetica en Alzheimer (EFIGA): 5R37AG015473, RF1AG015473, R56AG051876; the Health & Aging Brain Study – Health Disparities (HABS-HD), supported by the National Institute on Aging of the National Institutes of Health under Award Numbers R01AG054073, R01AG058533, R01AG070862, P41EB015922, and U19AG078109; the Korean Brain Aging Study for the Early Diagnosis and Prediction of Alzheimer’s disease (KBASE), which was supported by a grant from Ministry of Science, ICT and Future Planning (Grant No: NRF-2014M3C7A1046042); Memory & Aging Project at Knight Alzheimer’s

Disease Research Center (MAP at Knight ADRC): The Memory and Aging Project at the Knight-ADRC (Knight-ADRC). This work was supported by the National Institutes of Health (NIH) grants R01AG064614, R01AG044546, RF1AG053303, RF1AG058501, U01AG058922 and R01AG064877 to Carlos Cruchaga. The recruitment and clinical characterization of research participants at Washington University was supported by NIH grants P30AG066444, P01AG03991, and P01AG026276. Data collection and sharing for this project was supported by NIH grants RF1AG054080, P30AG066462, R01AG064614 and U01AG052410. We thank the contributors who collected samples used in this study, as well as patients and their families, whose help and participation made this work possible. This work was supported by access to equipment made possible by the Hope Center for Neurological Disorders, the Neurogenomics and Informatics Center (NGI: https://neurogenomics.wustl.edu/) and the Departments of Neurology and Psychiatry at Washington University School of Medicine; National Alzheimer’s Coordinating Center (NACC): The NACC database is funded by NIA/NIH Grant U24 AG072122. SCAN is a multi-institutional project that was funded as a U24 grant (AG067418) by the National Institute on Aging in May 2020. Data collected by SCAN and shared by NACC are contributed by the NIA-funded ADRCs as follows: P30 AG062429 (PI James Brewer, MD, PhD), P30 AG066468 (PI Oscar Lopez, MD), P30 AG062421 (PI Bradley Hyman, MD, PhD), P30 AG066509 (PI Thomas Grabowski, MD), P30 AG066514 (PI Mary Sano, PhD), P30 AG066530 (PI Helena Chui, MD), P30 AG066507 (PI Marilyn Albert, PhD), P30 AG066444 (PI John Morris, MD), P30 AG066518 (PI Jeffrey Kaye, MD), P30 AG066512 (PI Thomas Wisniewski, MD), P30 AG066462 (PI Scott Small, MD), P30 AG072979 (PI David Wolk, MD), P30 AG072972 (PI Charles DeCarli, MD), P30 AG072976 (PI Andrew Saykin, PsyD), P30 AG072975 (PI David Bennett, MD), P30 AG072978 (PI Neil Kowall, MD), P30 AG072977 (PI Robert Vassar, PhD), P30 AG066519 (PI Frank LaFerla, PhD), P30 AG062677 (PI Ronald Petersen, MD, PhD), P30 AG079280 (PI Eric Reiman, MD), P30 AG062422 (PI Gil Rabinovici, MD), P30 AG066511 (PI Allan Levey, MD, PhD), P30 AG072946 (PI Linda Van Eldik, PhD), P30 AG062715 (PI Sanjay Asthana, MD, FRCP), P30 AG072973 (PI Russell Swerdlow, MD), P30 AG066506 (PI Todd Golde, MD, PhD), P30 AG066508 (PI Stephen Strittmatter, MD, PhD), P30 AG066515 (PI Victor Henderson, MD, MS), P30 AG072947 (PI Suzanne Craft, PhD), P30 AG072931 (PI Henry Paulson, MD, PhD), P30 AG066546 (PI Sudha Seshadri, MD), P20 AG068024 (PI Erik Roberson, MD, PhD), P20 AG068053 (PI Justin Miller, PhD), P20 AG068077 (PI Gary Rosenberg, MD), P20 AG068082 (PI Angela Jefferson, PhD), P30 AG072958 (PI Heather Whitson, MD), P30 AG072959 (PI James Leverenz, MD); National Institute on Aging Alzheimer’s Disease Family Based Study (NIA-AD FBS): U24 AG056270; Religious Orders Study (ROS): P30AG10161,R01AG15819, R01AG42210; Memory and Aging Project (MAP - Rush): R01AG017917, R01AG42210; Minority Aging Research Study (MARS): R01AG22018, R01AG42210; the Texas Alzheimer’s Research and Care Consortium (TARCC), funded by the Darrell K Royal Texas Alzheimer’s Initiative, directed by the Texas Council on Alzheimer’s Disease and Related Disorders; Washington Heights/Inwood Columbia Aging Project (WHICAP): RF1 AG054023;and Wisconsin Registry for Alzheimer’s Prevention (WRAP): R01AG027161 and R01AG054047. Additional acknowledgments include the National Institute on Aging Genetics of Alzheimer’s Disease Data Storage Site (NIAGADS, U24AG041689) at the University of Pennsylvania, funded by NIA

