## Supplementary Figures for "Alzheimer’s Polygenic Risk Scores Are Not Interchangeable: Evidence from 1,752 Models"

### Supplementary Figure S1

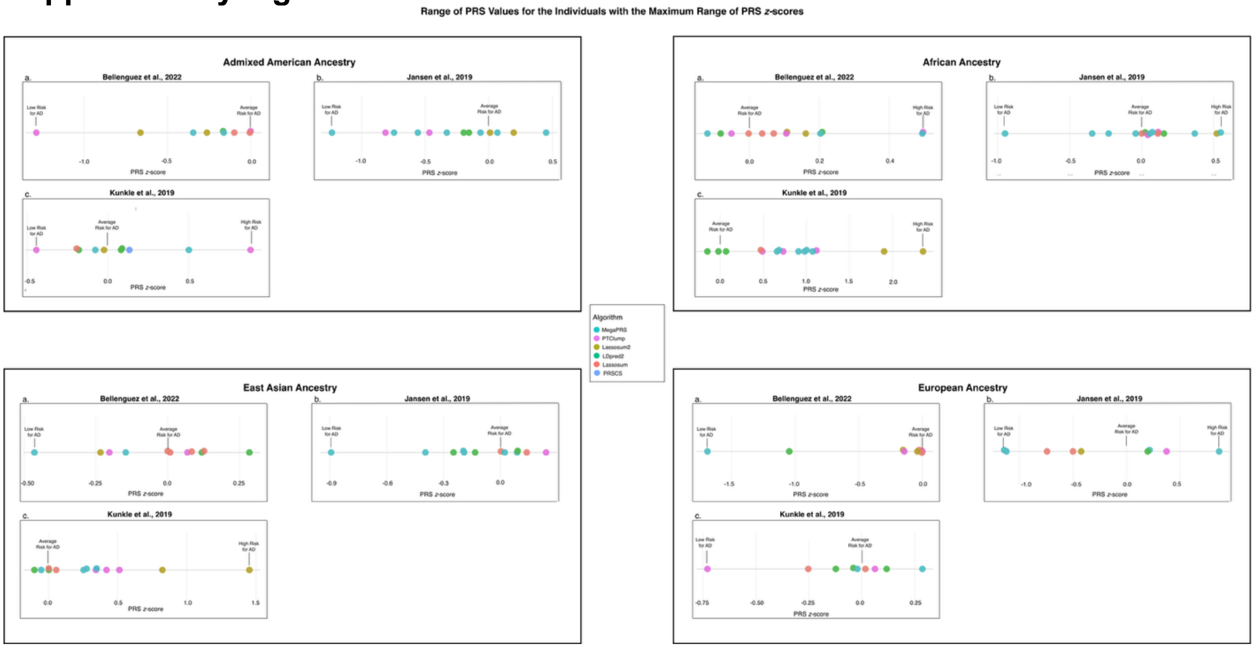

***Figure S1.*** *Range of uncorrelated (ρ < 0.3) PRS for AD calculated for the individual with the maximum range of PRS z-scores for each genetic ancestry (A: AMR, B: AFR, C: EAS, D: EUR) and then stratified by GWAS employed in the PRS calculation. PRS z-scores were computed using six different algorithms, indicated by color. Substantial variability in PRS values can arise for the same individual depending on the choice of algorithm and GWAS.*

### Supplementary Figure S2

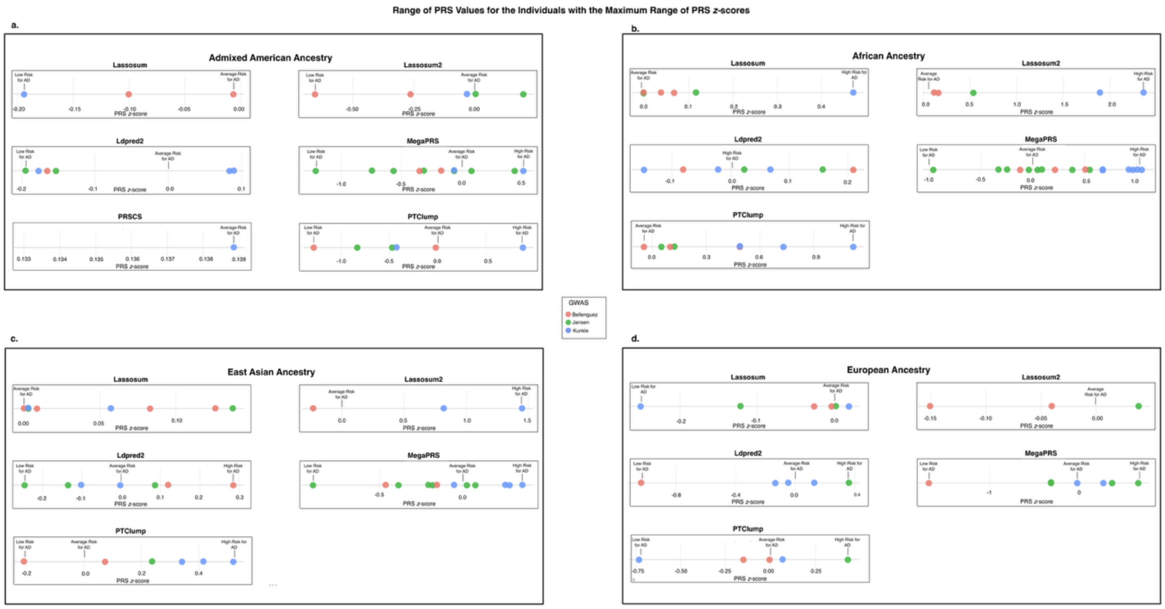

***Figure S2.*** *Range of uncorrelated (ρ < 0.3) PRS for AD calculated for the individual with the maximum range of PRS z-scores for each genetic ancestry (A: AMR, B: AFR, C: EAS, D: EUR) and then stratified by the algorithm employed in the PRS calculation. PRS z-scores were computed using three different GWAS summary statistics, indicated by color. Substantial variability in PRS values can arise for the same individual depending on the choice of algorithm and GWAS.*

### Supplementary Figure S3

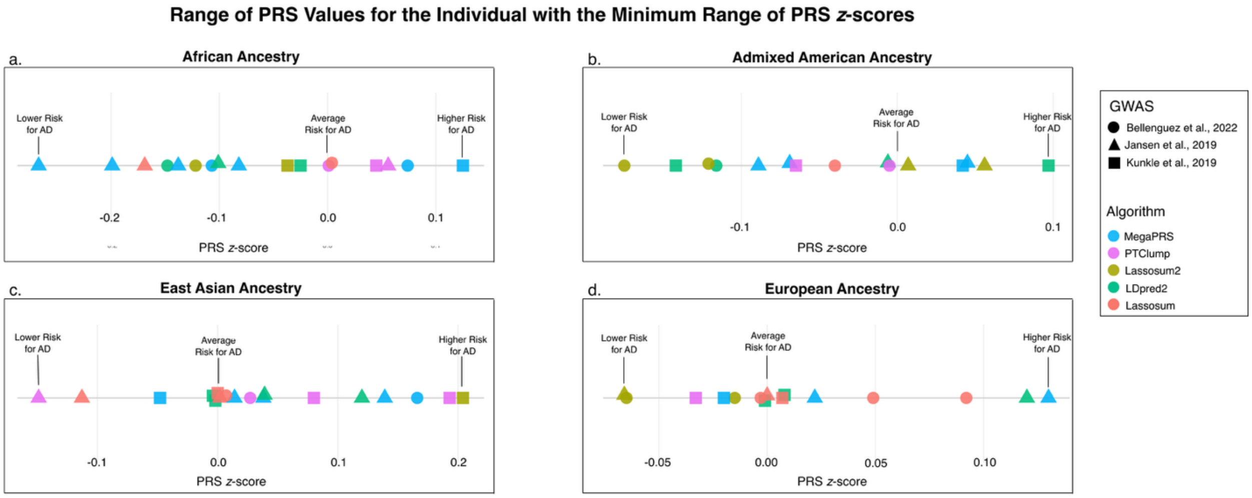

***Figure S3***. *Range of uncorrelated (ρ < 0.3) PRS for AD calculated for a single individual in each genetic ancestry (A: AFR, B: AMR, C: EAS, D: EUR). The individual with the minimum range of PRS z-scores, depending on the methodology, is shown. PRS z-scores were computed using five different algorithms, indicated by color, and three different GWAS summary statistics, indicated by shape. Vertical lines mark the low-risk, average risk, and high-risk thresholds for AD. The figure illustrates the substantial variability in PRS values that can arise for the same individual depending on the choice of algorithm and GWAS.*
